# Generalizability of Polygenic Risk Scores for Breast Cancer in the Multiethnic eMERGE Study

**DOI:** 10.1101/2020.08.17.20176685

**Authors:** Cong Liu, Nur Zeinomar, Wendy K. Chung, Krzysztof Kiryluk, Ali G. Ghravi, George Hripcsak, Katherine D. Crew, Ning Shang, Atlas Khan, David Fasel, Teri A. Manolio, Gail P. Jarvik, Robb Rowley, Ann E. Justice, Alanna K. Rahm, Stephanie M. Fullerton, Jordan W. Smoller, Eric B. Larson, Paul K. Crane, Ozan Dikilitas, Mary Beth Terry, Chunhua Weng

## Abstract

**Background:** The majority of polygenic risk scores (PRS) for breast cancer have been developed and validated using cohorts of European ancestry (EA). Less is known about the generalizability of these PRS in other ancestral groups.

**Methods:** The Electronic Medical Records and Genomics (eMERGE) network cohort dataset was used to evaluate the performance of seven previously developed PRS (three EA-based PRSs, and four non-EA based PRSs) in three major ancestral groups. Each PRS was separately evaluated in EA (cases: 3939; controls: 28840), African ancestry (AA) (cases: 121; controls: 1173) and self-reported LatinX ancestry (LA) (cases: 92; controls: 1363) women. We assessed the association between breast cancer risk and each PRS, adjusting forage, study site, breast cancer family history, and first three ancestry informative principal components.

**Results:** EA-based PRSs were significantly associated with breast cancer risk in EA women per one SD increase (odds ratio [OR] = 1.45, 95% confidence interval [CI] = 1.40–1.51), and LA women (OR = 1.41, 95% CI = 1.13–1.77), but not AA women (OR = 1.13, 95% CI = 0.92–1.40). There was no statistically significant association for the non-EA PRSs in all ancestry groups, LA including an LA-based PRS and an AA-based PRS.

**Conclusion:** We evaluated EA-derived PRS for estimating breast cancer risk using the eMERGE dataset and found they generalized well in LA women but not in AA women. For non-EA based PRSs, we did not replicate previously reported associations for the respective ancestries in the eMERGE cohort. Our results highlight the need to improve representation of diverse population groups, particularly AA women, in research cohorts.

## Introduction

Polygenic risk scores (PRS), which combine the effect of common genetic variants that individually confer minimal risk, have consistently shown the ability to stratify a woman’s risk of breast cancer[1]. Using large cohorts of European ancestry (EA) women, a breast cancer PRS In this study, we used the Electronic Medical Records and Genomics (eMERGE) network’s diverse data set to evaluate the performance of seven previously developed PRSs (three EA-based PRSs, and four non-EA based PRSs) for breast cancer in women of EA, African (AA) and LatinX (LA) ancestry. eMERGE is a network of academic medical centers in the United States developed in the Breast Cancer Association Consortium (BCAC), reported approximately 2-fold and 4-fold increases in breast cancer risk for women in the highest 10% and 1% of the PRS respectively, compared to women in the middle quantiles (40%-60%)[2]. This association has been replicated in validation studies using large cohorts of EA women, including the UK biobank and a Dutch prospective cohort [3, 4]. However, an important question remains about how PRSs perform in non-EA populations. Few studies have examined the performance of EA-derived PRSs in individuals of non-EA ancestry, and have reported either worse [5] or similar performance [6]. Further, the few non-EA based breast cancer PRSs that have been developed are constructed using datasets of substantially smaller sample sizes than EA-based PRS, and their performance has not been validated.

In this study, we used the Electronic Medical Records and Genomics (eMERGE) network’s diverse data set to evaluate the performance of seven previously developed PRSs (three EA-based PRSs, and four non-EA based PRSs) for breast cancer in women of EA, African (AA) and LatinX (LA) ancestry. eMERGE is a network of academic medical centers in the United States that has compiled electronic medical record (EMR) and genotype data. Understanding the performance of these PRS in diverse populations is crucial as we move towards clinical implementation of the PRS. As these PRS become validated and incorporated into clinical practice, they will also need to be integrated with other clinical covariates like family history to be able to apply the information for breast cancer risk stratification [7, 8]. With few exceptions[9], studies have not yet evaluated the performance of PRS using clinical data extracted from EMR. Our study aims to provide a systematic evaluation of the generalizability of breast cancer PRSs using the rich resources of the eMERGE network, including extensive breast cancer phenotyping algorithms, and a diverse population assembled across the networks’ federated environment.

## Methods

### Study Participants

The participants were women enrolled through the eMERGE network from eight medical centers with EMRs linked to genotype data. We identified breast cancer cases and controls through a validated EMR phenotyping algorithm (described below). We established ancestry by requiring the observed/self‐reported ancestry to match the genetic ancestry inferred by principal component analysis‐based k‐means group, as previously described [10]. Note for LA women, we only used self-report due to the diversity of admixture genetic background. We did not include Asian, Native American, and other ancestry groups given the small number of cases in the network for these ancestries. The institutional review board of each contributing institution approved the eMERGE study, and all participants provided written informed consent prior to study inclusion.

### PRS Models

We examined the performance of seven PRS models previously developed and tested in EA, AA, or LA women (**Table 1**). We reconstructed each PRS based on included variants and corresponding effect sizes in the original publications (details in **Supplementary Methods**). We included three PRS models developed in EA women, (BCAC-S, BCAC-L, UKBB), which included 313, 3820, and 5218 variants respectively [2, 11]. We also included two PRS models developed in or adapted to LA women (WHI-LA, 71 variants[5] and LATINAS, 179 variants[6]), as well as two PRS models developed in AA women (WHI-AA, 75 variants[5] and ROOT, 34 variants[12]) (**Table 1**). For EA women, we also evaluated subtype specific PRSs, specifically PRSs developed for estrogen receptor (ER)-positive and ER-negative breast cancers, as previously described [2]. We used PLINK 1.9 [13, 14] to calculate each PRS as a weighted sum using the –-score function (details in **Supplementary Methods**).

**Table 1:** Seven PRS models previously developed for the European ancestry or optimized for other ancestries.^a^ The number in the parenthesis represent the number of variants overlapped with the genotype dataset in eMERGE.^b^ Sample size of the original study, with the number of breast cancer cases presented in parenthesis.^c^ While the original publication states there are 180 variants in the PRS, one variant was removed due to low imputation quality, which left 179 variants in the supplementary table.

| PRS | Author (year),<br>Original study | # of variants<sup>a</sup><br>Original PRS (PRS<br>in eMERGE) | Sample size<sup>b</sup><br>Overall N (Breast cancer cases) | Ancestry |  |
| --- | --- | --- | --- | --- | --- |
|  |  |  |  | Derivation | Validation |
| **BCAC-S** | Mavaddat (2019) [2], Breast Cancer Association Consortium (BCAC) including 69 studies for development, and 10 prospective studies in the test set. Also validated in the UK biobank. | 313 (193) | Development:<br>169,092 (94,075)<br>Test set: 29,751 (11,428) | European | European |
| **BCAC-L** | Same as BCAC-S | 3820 (2213) | Development:<br>169,092 (94,075)<br>Test set: 29,751 (11,428) | European | European |
| **LATINAS** | Shieh (2020) [6], Pooled case-control analysis of eight studies of US Latinas and Latin American women (from Mexico, Colombia, and Peru). Ancestry was predominantly European and Indigenous American. | 179<sup>c</sup> (129) | 12,280 (4,658) | European | LatinX |
| **ROOT** | Wang (2018) [12], The Root consortium: breast cancer GWAS in the African diaspora including participants of African ancestry from Nigeria, USA, and Barbados. | 34 (30) | 3,686 (1,657) | African | African |
| **UKBB** | Khera (2018) [11], UK biobank cohort phase 1 and phase 2 genotype data release. | 5218 (2768) | 190,040 (3,215) | European | European |
| **WHI-AA** | Allman (2015) [5], Self-reported African American women from the Women's Health Initiative (WHI) SNP Health Association Resource (SHARe). | 75 (67) | 7,539 (421) | African | African |
| **WHI-LA** | Allman (2015) [5], Self-reported Hispanic women in the WHI SHARe. | 71 (64) | 3,363 (147) | LatinX | LatinX |

### Phenotypes and Genotypes

Details of the eMERGE genotyping procedures and quality control procedures have been previously described [10] and are in **Supplementary Methods**. We used EMR data to phenotype each participant, including breast cancer case-control status, demographic information, ER status, family history, and age (details in the **Supplementary Methods)**. We classified women as breast cancer cases or controls using a validated phenotyping algorithm (>95% positive predictive value for both cases and controls) that incorporated information from the ICD-9/ICD-10 diagnostic codes, breast pathology reports, and medication lists. The breast cancer phenotyping workflow is shown in **Supplementary Figure S1**.

### Statistical Analysis

To evaluate the performance of each PRS, we standardized the PRSs to have a risk score unit expressed as a standard deviation (SD) of the control distribution. The association of the standardized PRSs and breast cancer risk was evaluated by logistic regression adjusted for the first three ancestry-specific principal components [10], age, family history of breast cancer, and study site. We defined age as the difference between the year the phenotyping algorithm was executed (*i.e*., 2019) and year of birth. In addition, we examined the association of breast cancer by percentiles of PRS, compared to the middle quantile (40–60%), or to the remainder of the population. We also used a Cox proportional hazards regression model with the same covariates to assess the association of breast cancer risk when the outcomes were time to the breast cancer events. Women were followed starting from first known date in the EMR to either diagnosis of breast cancer (defined as the age of the first breast cancer-related ICD codes in the EMR), or age at last observation in the EMR.

To examine the discrimination of each PRS, we estimated the area under the receiver operator characteristic curves (AUC), with only the PRS used as a predictor. To estimate the percentage of the total variance in breast cancer risk explained by PRS, we used Nagelkerke’s pseudo R^2^ from logistic regression models. We also chose the PRS that was the most strongly associated with breast cancer within each ancestry (UKBB in EA and AA women, and BCAC-L model in LA) to estimate the cumulative risk of breast cancer for high PRS risk (> average PRS + 2 SD), moderate PRS risk (> average PRS + 1 SD) and population risk (within 0.5 SD of average PRS) individuals in each ancestry using iCARE [15] (details in **Supplementary Methods**).

We assessed statistical power for testing associations of PRSs with breast cancer given sample size for each ancestry. Based on ancestry-specific empirical effect sizes of the PRS obtained from the literature, we assumed odds ratios (ORs) of 1.61 [2], 1.23 [5] and 1.58 [6] for EA, AA and LA women, respectively. Power analysis was then conducted for each ancestry by using R function pwr.f2.test(). Our power analysis shows we have 100%, 24% and 94% power to detect an association of above assumed ORs for EA, AA, and LA respectively. When we assumed a moderate PRS effect size (OR = 1.39) for LA women as reported in Allman *et al*. [5], we observed 66% power to detect an association in LA women. All analyses were conducted in R v.3.0.2. All statistical tests were two sided, and P-values < 0.05 were considered significant.

## Results

After applying the breast cancer phenotyping algorithm, our cohort included 35527 women, including 32779 EA, 1293 AA, and 1455 LA women **(Table 2)**. The total number of variants included in the PRS calculation for each model is presented in **Table 1**. On average, 73% of the variants included in the original PRS overlapped with the eMERGE genotype dataset.

**Table 2:** Participant Characteristics. EA = European ancestry; AA = African ancestry; LA = LatinX ancestry^a^ Age was calculated at the time of electronic phenotyping algorithm deployment. ^b^ Age at breast cancer diagnosis was defined as the age at the first breast cancer ICD related code.

| **Characteristic** | **EA**<br>**(n = 32779)** | **AA**<br>**(n=1293)** | **LA**<br>**(n=1455)** |
| --- | --- | --- | --- |
| **Age, years (mean ± SD)<sup>a</sup>** | 65.9 ± 17.7 | 53.7 ± 18.0 | 56.4 ± 19.0 |
| **Breast cancer diagnosis, n (%)** | 3939 (12.0) | 121 (9.4) | 92 (6.3) |
| **Age at breast cancer diagnosis<sup>b</sup>, years**<br>**(mean ± SD)** | 60.5 ± 13.1 | 55.9 ± 14.8 | 54.1 ± 12.7 |
| **Estrogen receptor status, n (% of cases)** |  |  |  |
| Positive | 1052 (26.7) | 20 (16.5) | 22 (23.9) |
| Negative | 241 (6.1) | 15 (12.4) | 4 (4.3) |
| Missing | 2646 (67.2) | 86 (71.1) | 66 (71.7) |
| **eMERGE site, n (%)** |  |  |  |
| Columbia University Medical Center | 203 (0.6) | 73 (5.6) | 160 (11) |
| Geisinger | 1331 (4.1) | 4 (0.3) | 8 (0.5) |
| Partners Healthcare | 13416 (40.9) | 926 (71.6) | 1096 (75.3) |
| Kaiser Permanente Washington Health<br>Research Institute / University of Washington | 1663 (5.1) | 66 (5.1) | 55 (3.8) |
| Marshfield Clinic Research Foundation | 2837 (8.7) | 0 | 8 (0.5) |
| Mayo Clinic | 3542 (10.8) | 9 (0.7) | 22 (1.5) |
| Northwestern University | 1900 (5.8) | 215 (16.6) | 23 (1.6) |
| Vanderbilt University | 7887 (24.1) | 0 | 83 (5.7) |

### Association of PRS with breast cancer risk in women of European ancestry (EA)

Our primary analysis examined the association of BCAC-S, BCAC-L, and UKBB in 3939 breast cancer cases and 28840 control EA women and is shown in **Figure 1A**. We found significant associations with overall breast cancer risk for all three PRSs examined; (BCAC-L OR: 1.40, 95% confidence interval (CI): 1.35–1.45; BCAC-S OR: 1.35, 95% CI: 1.30–1.40; UKBB OR: 1.45, 95% CI: 1.40–1.51). The hazard ratios (HRs) for breast cancer per one SD higher PRS are 1.06 (95% CI: 1.02–1.09), 1.07 (95% CI: 1.04–1.10) and 1.09 (1.06–1.13) for the BCAC-L, BCAC-S, and UKBB, respectively.

**Figure 1:**
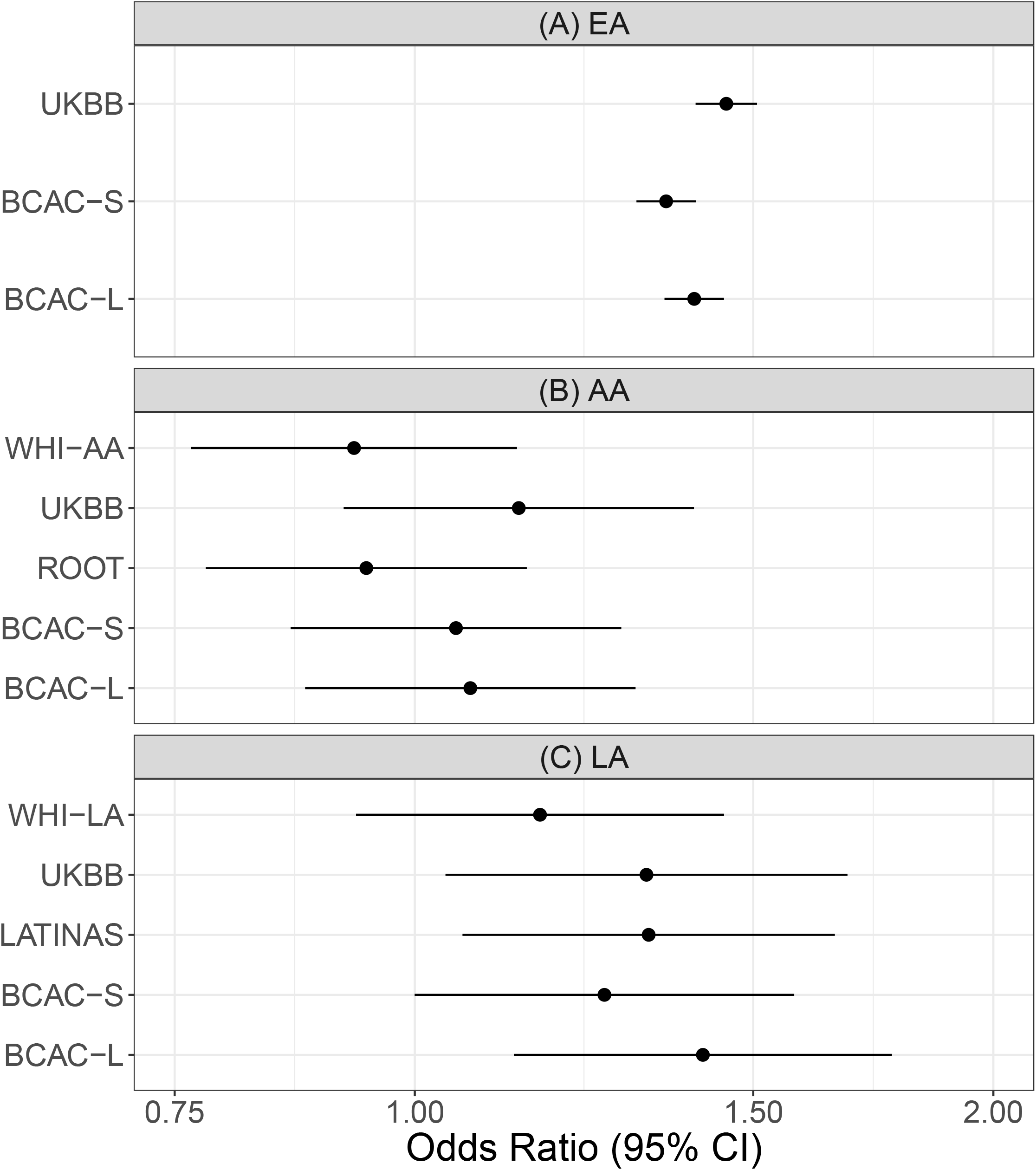
The association of the different PRSs and breast cancer risk in women of (A) European ancestry (EA); (B) African ancestry (AA); and (C) LatinX ancestry (LA) in the eMERGE cohorts. The odds ratios and 95% confidence intervals of breast cancer per standard PRS unit increase, adjusted for the first three ancestry-specific principal components, age, family history, and study site are shown. Breast cancer cases and controls are defined according to breast cancer phenotyping algorithm. BCAC-S includes 313 variants in the original PRS, BCAC-L includes 3820 variants in the original PRS, WHI-LA includes 71 variants in the original PRS and was optimized for LA women, WHI-AA includes 75 variants in the original PRS and was optimized for AA women, UKBB includes 5218 variants in the original PRS, ROOT includes 34 variants in the original PRS and was optimized to AA women, and LATINAS includes 179 variants in the original PRS and was optimized for LA women.

As illustrated in **Figure 2**, this association with breast cancer risk was largest in the extremes of the PRS distribution, with ORs ranging from 2.31–2.97 for the three different PRSs examined for those in the highest 1% of the PRS compared to those in the middle quantile. For example, for the UKBB PRS, compared to women in the middle quantile (40–60%), we observed a nearly 3- fold increase in risk for women in the top 1% (OR: 2.97; 95% CI: 2.23 – 3.94) (**Figure 2**). We observed significant association and similar effect sizes for each of the three PRS with overall breast cancer when we compared the extreme ends of the distribution to those in the remainder of the PRS distribution. For example, we observed an OR of 2.66 (95% CI: 2.03–3.48) for the top 1% of the UKBB PRS distribution, compared to the women in the remaining 99% of the distribution (**Supplementary Figure S2**). We found similar AUCs for each of the three PRSs examined. The AUC for the BCAC-L, BCAC-S, and UKBB PRS in EA women was 0.60 (95% CI:0.59–0.61), 0.59 (95% CI:0.58–0.60), and 0.61 (95% CI:0.60–0.62), respectively (**Table 3**). Variance for breast cancer risk as quantified by Nagelkerke’s R^2^ value was 3.0% for the best performing model from UKBB.

**Table 3:** The AUC (95% CI) for breast cancer prediction using PRS as a single predictor in women of different ancestries. The Nagelkerke’s R^2^ (%) of a fitted logistic regression with PRS as a single variable is calculated.

| PRS | EA<br>(3939 breast cancer cases/<br>28840 controls) |  | AA<br>(121 breast cancer cases/<br>1173 controls) |  | LA<br>(92 breast cancer cases/<br>1363 controls) |  |
| --- | --- | --- | --- | --- | --- | --- |
|  | AUC (95% CI) | R<sup>2</sup> | AUC (95% CI) | R<sup>2</sup> | AUC (95% CI) | R<sup>2</sup> |
| BCAC-L | 0.60 (0.59-0.61) | 2.54 | 0.52 (0.47-0.57) | 0.07 | 0.57 (0.5-0.63) | 1.14 |
| BCAC-S | 0.59 (0.58-0.60) | 2.04 | 0.51 (0.46-0.56) | 0.01 | 0.54 (0.48-0.60) | 0.41 |
| UKBB | 0.61 (0.60-0.62) | 3.00 | 0.54 (0.49-0.60) | 0.27 | 0.54 (0.47-0.60) | 0.27 |
| LATINAS | - | - | - | - | 0.57 (0.50-0.63) | 0.90 |
| ROOT | - | - | 0.49 (0.43-0.55) | 0.67 | - | - |
| WHI-AA | - | - | 0.48 (0.42-0.54) | 1.55 | - | - |
| WHI-LA | - | - | - | - | 0.53 (0.47-0.59) | 1.60 |

**Figure 2:**
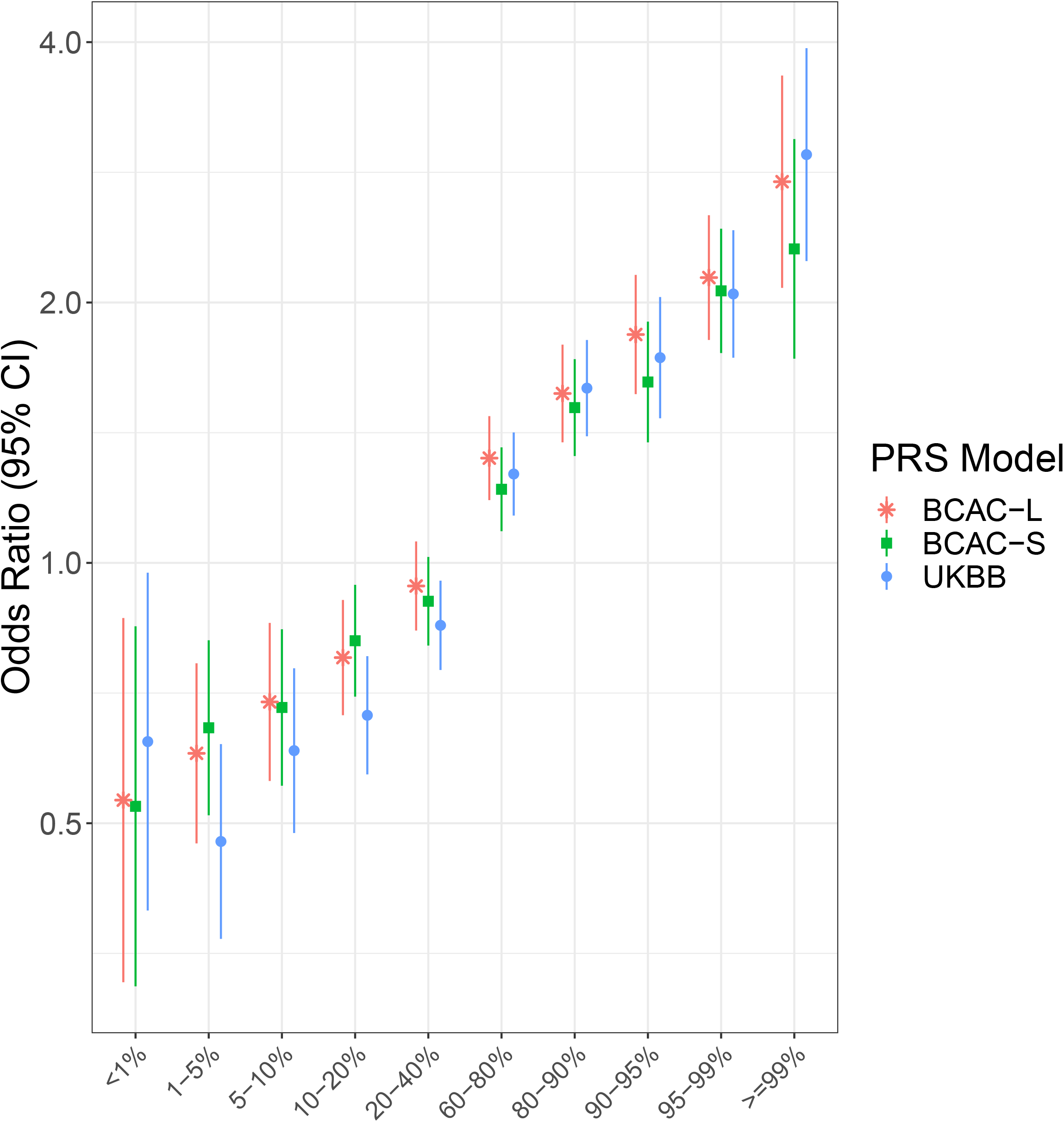
The association of the PRSs (for different quantities relative to the middle quantile, 40–60%) and overall breast cancer risk in women of European ancestry. The odds ratio and 95% confidence intervals, adjusted for the first three ancestry-specific principal components, age, family history, and study site are shown. BCAC-S includes 313 variants in the original PRS, BCAC-L includes 3820 variants in the original PRS, and UKBB includes 5218 variants in the original PRS.

When we examined the association of PRS by ER status (ER-positive and ER-negative), we found significant associations for both ER–positive and ER-negative breast cancers, although the observed effect size was larger for ER-positive breast cancers (**Figure 3**). The findings were nearly identical for both overall PRSs and subtype-optimized PRSs. For example, when we examined the BCAC-S PRS developed for overall BC, the OR per SD of the PRS for ER-negative breast cancer was 1.17 (95% CI:1.03–1.33) and 1.50 (95% CI:1.4–1.59) for ER-positive breast cancer, and these findings were identical to subtype optimized PRSs, BCAC-S-H-ERN and BCAC-S-H-ERP.

**Figure 3:**
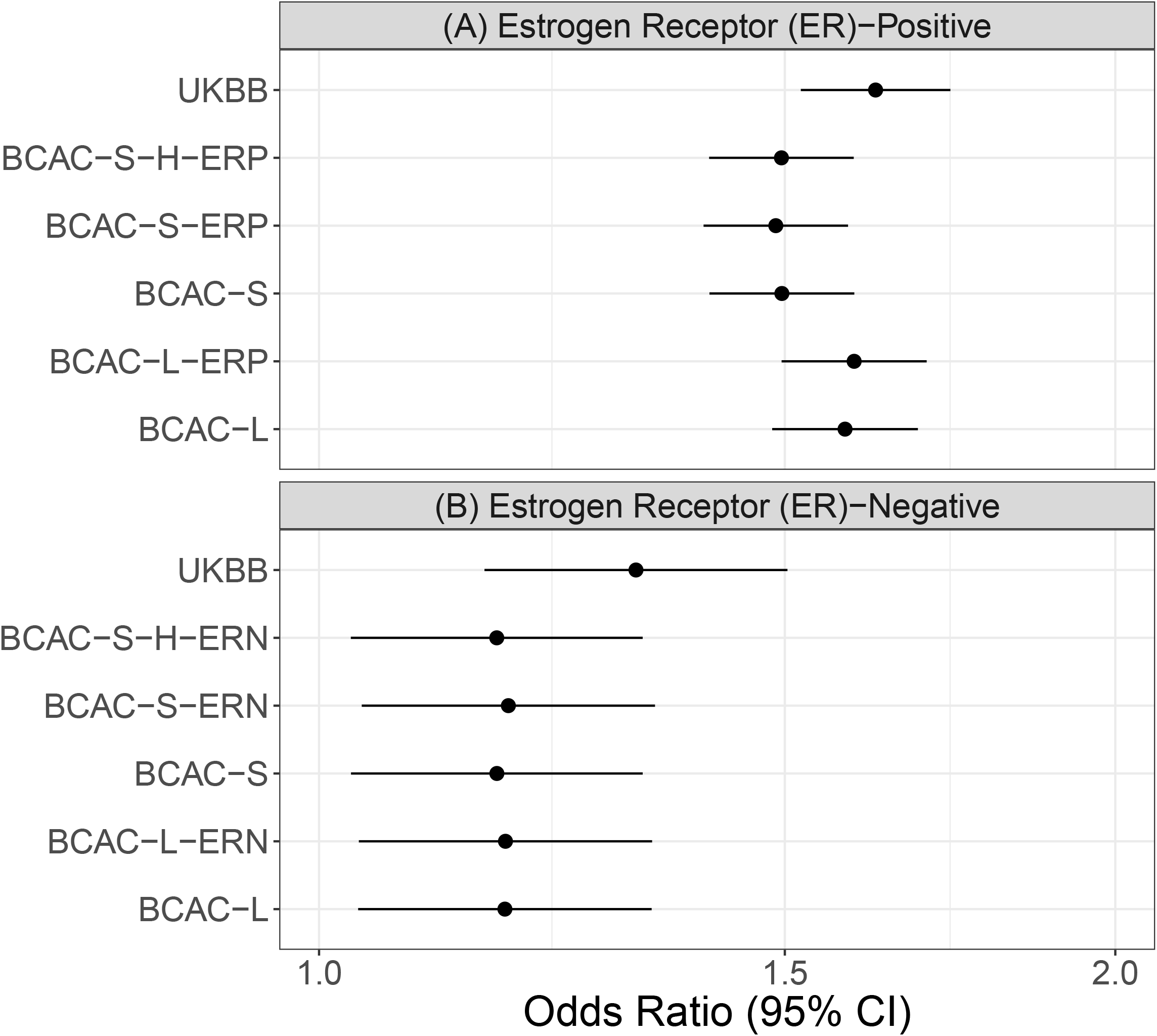
The association between different PRSs and (A) estrogen receptor (ER)-positive and (B) ER-negative breast cancer for women of European ancestry women. Odds ratios and 95% confidence intervals are presented for each of the of breast cancer subtypes (A: ER-positive and B: ER-negative) per standard PRS unit increase.

### Association of PRS with breast cancer risk in women of African ancestry (AA)

In 121 breast cancer cases and 1173 control AA women, we examined the association of five previously developed PRSs: three developed and tested in EA women (BCAC-S, BCAC-L, and UKBB) and two developed in AA women (ROOT and WHI-AA). We did not observe a significant association between any of these PRSs and breast cancer risk (**Figure 1B**). The largest effect size we observed in AA women was for UKBB, with an OR per SD of the PRS of 1.13, although not statistically significant. Compared to EA women, we observed lower AUCs for each of the three PRSs in AA women (BCAC- L: 0.52 (95% CI: 0.47, 0.57); BCAC-S: 0.51 (95% CI:0.46–0.56); and UKBB: 0.54 (95% CI:0.49–0.60) (**Table 3**). The AUC for the PRSs developed in AA women was 0.49 for ROOT and 0.48 for WHI-AA.

### Association of PRS with breast cancer risk in LatinX women (LA)

We examined the association of five PRSs (BCAC-S, BCAC-L, UKBB, WHI-LA, and LATINAS), two of which were developed in or adapted to LA (WHI-LA and LATINAS) in 92 breast cancer cases and 1363 LA controls. For LA women, we observed a significant association for overall breast cancer risk for four of the PRSs examined (BCAC-L, UKBB, LATINAS, BCAC-S), with ORs per SD of the PRS ranging from 1.26–1.41 (**Figure 1C**). BCAC-L PRS had an OR similar to that in EA women (BCAC-L OR in LA women: 1.41, 95% CI: 1.13–1.77). When we examined the association of PRS quantiles, we observed more pronounced associations for those at the higher end of the PRS risk (≥ 80%) compared to those in the middle quantiles(40–60%) for BCAC-L, LATINAS, and WHI-LA; however, none of these associations were significant, likely due to small numbers in these strata. For example, women in the top 20% of the BCAC-L PRS distribution had a two-fold increase in breast cancer risk compared to women in the middle quantiles (OR: 2.05, 95% CI: 0.97–4.30). Compared to EA women, we found lower AUCs in LA women for BCAC-L, BCAC-S and UKBB, and we observed the highest AUCs for the BCAC-L and LATINAS PRSs (AUC: 0.57, 95% CI: 0.5–0.63) in LA women (**Table 3**).

### Estimation of absolute risk of breast cancer

As shown in **Figure 4**, there were differences in cumulative absolute breast cancer risk by categories of PRS for EA, AA, and LA women. When we compared those in the highest PRS risk category to those at population risk, EA women had larger risk gradient than AA and LA women. For example, EA, AA, and LA women in the lowest PRS risk category had a cumulative breast cancer risk of 12.6%, 11.9% and 9.2%, respectively, by age 80 years, whereas women in the highest PRS risk category had 28.2%, 15.6% and 20.0% cumulative risk, respectively (**Figure 4**).

**Figure 4:**
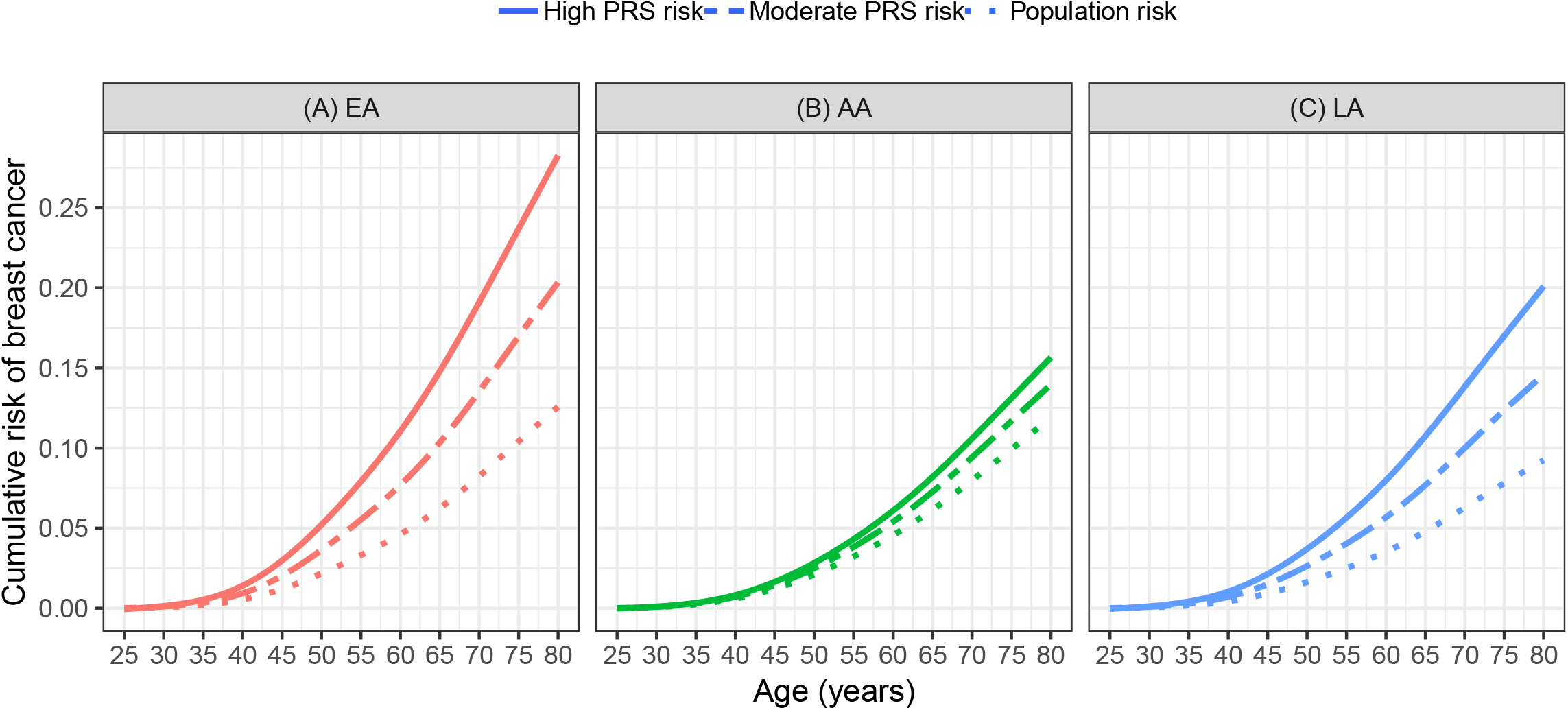
Cumulative risk (from birth) of breast cancer predicted by UKBB PRS model in women of European (EA) and African (AA) ancestry, and BCAC-L PRS model in LatinX women (LA)

## Discussion

For PRSs developed in cohorts of EA women (UKBB, BCAC-L, BCAC-S), we replicated significant association for increased breast cancer risk in EA women, although the ORs observed in our study were smaller in magnitude than the original studies (**Supplementary Table S4**). For example, BCAC-L had an OR of 1.40 in EA women compared to an OR of 1.66 reported in the original study [2]. Moreover, similar to other studies investigating the generalizability of PRSs in EA and non-EA cohorts, we found EA-based PRSs generalized well in LA women, but not in AA women. This is likely due to LA individuals in the US having a greater proportion of European ancestry than AA individuals [16]. Given that the majority of the participants in breast cancer-related GWAS are EA women, the lack of significant association of EA-based PRSs with breast cancer risk in AA women in our study is not surprising and is consistent with the PRS performance in non-EA cohorts for other diseases [17–21]. Worthy of note is the limited sample size for LA and AA women in our study, particularly for AA women where we had limited power. The poor generalizability can also be partly explained by differences in risk allele frequencies and LD patterns among diverse ancestries [21, 22]. This PRS coupled with other risk factors may influence clinical recommendations at an individual level such as enhanced screening. For example, the predicted cumulative risk by age 80 years for EA women in the lowest category of PRS is similar to the population risk (12.5%) and increases to 27% for EA women in the highest PRS category.

For PRSs developed in non-EA study populations (WHI-AA, WHI-LA, and ROOT) or adapted to non-EA population (LATINAS), we did not replicate the previously reported associations in the eMERGE cohort for either LA or AA women, except for the LATINAS in LA women. LATINAS is a multi-ethnic PRS that used effect sizes obtained from the EA population and further developed the PRS in a cohort of LA women, suggesting that combining training data from EA samples could improve the observed associations in non-EA populations [23–25].

Because the PRSs developed in non-EA studies are often based on much smaller GWAS cohorts, the uncertainty of the effect sizes used in those PRSs is larger, making their predictive power lower in the non-EA population [6, 12]. A recent study showed genome-wide PRSs with a larger number of included variants were more strongly associated with CHD than restricted PRSs [26]. The non-EA based PRSs included fewer variants due to smaller sample size in the discovery GWAS cohort, possibly additionally contributing to their weaker generalizability. Furthermore, even with an adequate sample size for non-EA populations, limitations inherent to the genotyping platforms used in GWAS [19] can make this subpopulation optimization theoretically insufficient to reduce the bias if the sub-population risk allele is not captured by the genotype platform, which is possible as many array designs are based on EA samples. Moreover, AA women’s 40% higher mortality rate [27], often attributed to later stage of diagnosis and related preventative healthcare barriers, create the needs to increase diversity in genomic studies so that future clinical applications of the PRS do not exacerbate existing health disparities.

The eMERGE [28] and the All of Us Research Program [29] are two programs actively involved in increasing their recruitment of diverse patients to help address the gap. A key aspect of these programs is derived from EMRs, providing a scalable approach to independently validate previously developed PRSs for different phenotypes in multiple clinical operation sites [30–33].

We found similar magnitude of PRS association in EA women across all study sites, except for Vanderbilt University (**Supplementary Figure S3**). This difference might be caused by the heterogeneity in the genotyping platforms and/or EMR systems [34, 35]. Continued efforts are required to address heterogeneity in the genotyping platforms and advance the unification of phenotypic information extracted from EMRs [36–40]. Our sensitivity analysis suggested that defining cases and controls based on a validated phenotyping algorithm achieved a slightly stronger association than the cohort when case-control definition was solely based on ICD codes (**Supplementary Table S5**). By utilizing EMR data elements such as drugs prescribed and ER-status, our phenotyping algorithm enabled analyses of disease subtypes. However, implementing a phenotyping algorithm is more labor-intensive than simply using ICD-codes and continued efforts will be required to develop portable and computable phenotypes [39, 41]. In addition, other data challenges such as incomplete records or inconsistent documentation [42, 43] should be addressed while using the EMR for translational study especially in time to event analysis.

In summary, we found EA-based PRSs were significantly associated with breast cancer risk in EA women in the eMERGE network. We also found that these EA-based PRSs generalized well to LA women but not to AA women. Additionally, we found that PRS developed on small, non-EA GWAS studies did not generalize well in the respective ancestry group. Our results highlight the need to increase the participation of racially and ethnically diverse patients, especially AA women, in research cohorts, and suggest that until well-developed and validated PRSs for non-EA women become available, the current PRSs developed based on EA cohorts could be adapted for LA women, but not AA women in clinical settings.

## Data Availability

Data are available from the authors upon reasonable request and with permission of eMERGE network

## Acknowledgments

We would like to thank all the investigators and participants of the electronic Medical Records and Genomics (eMERGE) Network. The eMERGE Network was initiated and funded by National Human Genome Research Institute (NHGRI) through the following grants: U01HG006828 (Cincinnati Children’s Hospital Medical Center and Boston Children’s Hospital); U01HG006830 (Children’s Hospital of Philadelphia); U01HG006389 (Essentia Institute of Rural Health, Marshfield Clinic Research Foundation, and Pennsylvania State University); U01HG006382 (Geisinger Clinic); U01HG006375 (Group Health Cooperative and the University of Washington); U01HG006379 (Mayo Clinic); U01HG006380 (Icahn School of Medicine at Mount Sinai); U01HG006388 (Northwestern University); U01HG006378 (Vanderbilt University Medical Center); and U01HG006385 (Vanderbilt University Medical Center serving as the Coordinating Center). This phase of the eMERGE network was initiated and funded by the NHGRI through the following grants: U01HG8657 (Group Health Cooperative/University of Washington); U01HG8685 (Brigham and Women’s Hospital); U01HG8672 (Vanderbilt University Medical Center); U01HG6379 (Mayo Clinic); U01HG8679 (Geisinger Clinic); U01HG8680 (Columbia University Health Sciences); U01HG8684 (Children’s Hospital of Philadelphia); U01HG8673 (Northwestern University); U01HG8701 (Vanderbilt University Medical Center serving as the Coordinating Center); U01HG8676 (Partners Healthcare and the Broad Institute); U54MD007593 (Meharry Translational Research Center); and U01HG8664 (Baylor College of Medicine). CL was additional supported by National Library of Medicine/National Human Genomic Research Institute Grant R01LM012895. NZ was supported by the National Institutes of Health (NIH) National Center for Advancing Translational Sciences (NCATS), TL1 Training Program TL1TR001875. The contents of this article are solely the responsibility of the authors and do not necessarily represent the official views of the National Institutes of Health.

## Notes

Authors have no conflicts of interest to declare

